# Endogenous retroelement activation is implicated in IFN-α production and anti-CCP autoantibody generation in early RA

**DOI:** 10.1101/2024.01.17.24301287

**Authors:** Faye AH Cooles, Gemma V Pedrola, Najib Naamane, Arthur G Pratt, Ben Barron-Millar, Amy E Anderson, Catharien MU Hilkens, John Casement, Vincent Bondet, Darragh Duffy, Fan Zhang, Ruchi Shukla, John D Isaacs

## Abstract

**Objectives:** Endogenous retroelements (EREs) stimulate type 1 interferon (IFN-I) production but have not been explored as potential interferonogenic triggers in Rheumatoid Arthritis (RA). We investigated ERE expression in early RA (eRA), a period where IFN-I is increased.

**Methods:** ERE expression in DMARD naïve eRA whole blood (LINE1; RT-PCR) and bulk synovial tissue (LTR5, LINE1, SINE; Nanostring) was examined alongside IFN-α activity. Circulating lymphocyte subsets, including B cell subsets, from eRA patients and early psoriatic arthritis (PsA), were flow cytometrically sorted and similarly examined. Existing established RA and osteoarthritis (OA) synovial single-cell sequencing data was re-interrogated to identify repeat elements, and associations explored.

**Results:** There was significant co-expression of all ERE classes and *IFNA* in eRA synovial tissue (n=22, p<0.0001) and significant positive associations between whole blood LINE1 expression (n=56) and circulating IFN-α protein (p=0.018) and anti-CCP titres (p<0.0001). ERE expression was highest in circulating eRA B cells, particularly naïve B cells compared with PsA, with ERE regulation by SAMDH1 implicated and associations with *IFNA* again observed. Finally, in established RA synovium, LTRs, particularly ERVK, were most increased in RA compared with OA where, for all synovial subsets (monocytes, B cells, T cells and fibroblasts), ERE expression associated with increased IFN-I signalling (p<0.001).

**Conclusions:** Peripheral blood and synovial ERE expression is examined for the first time in eRA highlighting both a potential causal relationship between ERE and IFN-I production and an intriguing association with anti-CCP autoantibodies. This suggests EREs may contribute to RA pathophysiology with implications for future novel therapeutic strategies.

## Introduction

Type 1 interferons (IFN-I) have pleiotropic effects on the immune system and prime cellular responses to effectively clear, typically viral, infection (1). In this context widespread cellular activation is desirable but in the absence of infection, IFN-I associated increased cellular priming/activation can be inappropriate (2). Excess IFN-α can promote breach of tolerance in autoantibody producing B cells (3, 4) as well as facilitating more effective presentation of antigen, potentially of self-components (5, 6). We have previously demonstrated increased IFN-I signaling and serum IFN-α levels in early rheumatoid arthritis (RA) with negative prognostic implications on initial disease control and clinical outcomes (7, 8). An elevated interferon gene signature (IGS) also increases the likelihood of progression to RA in at risk populations, such as those with anti-cyclic citrullinated peptide (CCP) positive arthralgia (7–13). However, it remains unknown what drives this IFN-I release in early RA.

Endogenous retrotransposons or retroelements (EREs) are sequences of DNA derived from ancient transposable elements, such as retroviruses, that have been historically incorporated into the genome (14). While the majority are inactive, some have retained transcriptional activity and their replication cycle and organization is similar to exogenous retroviruses, such as human immunodeficiency virus (HIV) (15). They can be sub-divided into endogenous retroviruses (ERVs), often detected as long terminal repeats (LTRs), long interspersed nuclear element 1 (LINE-1) and short interspersed nuclear elements (SINEs), most commonly “Alu”. Retrotransposons can replicate, generating a strand of mRNA and, subsequently, a double stranded RNA product which then inserts into a unique region of the genome, often separate from the area of origin; depending upon the site of insertion, this potentially disrupts protein coding regions (5, 16). This process of active retrotransposition results in the accumulation of cytosolic single stranded DNA, which triggers an IRF3-dependent innate immune response, including release of IFN-I. (17–19). Indeed, single mutations in human genes that regulate retroelement replication, such as *TREX1* or *SAMHD1*, cause type 1 interferonopathies such as Aicardi-Goutieres syndrome (AGS) (20).

The potential for an association between EREs and IFN-I production in autoimmunity is increasingly appreciated (17–21). In diseases where IFN-I are known to play a pathogenic role, such as systemic lupus erythematous (SLE) or primary Sjogren’s syndrome (PSS), there is evidence of increased ERE activity in disease relevant tissue associated with increased local IFN-α production (21). Established RA synovium was shown to overexpress LINE1 nearly 2 decades ago (22, 23). Some ERVs retain their ability to produce viral protein, and ERV viral protein products have been detected in the peripheral circulation of RA patients and linked to autoantibody generation (24–27). However, EREs have not been examined in RA in relation to IFN-I production nor in early RA, a period where IFN-I signaling, and autoantibody generation, is important. We therefore explored ERE expression in whole blood, circulating lymphocyte subsets and synovial tissue from treatment naïve early RA patients, examining associations with IFN-I generation and autoantibody status.

## Methods

### Patient cohorts

Glucocorticoid and disease-modifying anti-rheumatic drug (DMARD)-naïve patients attending Newcastle-upon-Tyne Hospitals, were enrolled for this study from the Northeast Early Arthritis Cohort (NEAC) if their initial consultant rheumatologist diagnosis was RA (with reference to 2010 ACR/EULAR RA classification criteria; early RA patents, eRA) or psoriatic arthritis (PsA, determined an early disease control group). Contemporaneous clinical parameters were recorded, including disease activity scores (DAS-28-ESR), immunoglobulins (IgG, IgA, IgM), inflammatory markers (CRP and ESR) and serological status, (rheumatoid factor and anti-cyclic citrullinated peptide titres; RF and anti-CCP).

### Patient and public involvement

Patients and the public were not formally involved in the design, conduct or reporting of this research.

### Interferon gene signature (IGS) and serum cytokines

Serum was spun and frozen within 4 hours of blood draw, undergoing no more than one freeze-thaw cycle before measurement of IFN-γ, IL-6, IL-12p70, TNF-α, IL-1β, IL-2, IL-13, IL-4, IL-10 by MSD technology (Meso Scale Discovery, MD, USA) as per manufacturers’ instructions. Serum IFN-α was measured using the digital Simoa platform as described (7). Serum IFN monoclonal antibodies (specific for all IFN-α subtypes) were isolated from APECED patients (28) and provided to DD by Immunoqure under an MTA. The IGS (interferon gene signature) was generated from whole blood RNA as described previously (29). The mean expression of 5 interferon response genes (IRGs) *MxA, IFI6, OAS1, ISG15* and *IFI44L* was termed the IGS as previously described (8).

### Flow cytometric cell sorting

For all samples peripheral blood mononuclear cells (PBMCs) were isolated from whole blood using density centrifugation and underwent immediate flow cytometric sorting. Plasmacytoid dendritic cells (pDCs), conventional CD1c^+^ DCs, CD4^+^ T cells, CD8^+^ T cells, CD19^+^ B cells, and CD14^+^ monocytes were sorted as previously described (29) and B cell subsets including naïve B cells (CD19^+^IgD^+^CD27^−^), memory B cells (CD19^+^IgD^−^CD27^+^), CD5^+^ B cells (CD19^+^CD5^+^) and age associated B-cells (ABCs, CD19^+^CD11c^+^CD21^−^) were flow cytometrically sorted from both early RA and early PsA patient PBMCs as previously described (30).

### Endogenous retroelement quantification

#### Whole blood and circulating lymphocytes

Whole blood RNA was isolated using the Tempus Spin Isolation Kit (Tempus, Thermo-Fisher Scientific, MA, USA) and treated with TurboDNase (Ambion, TX, USA) to remove any contaminating genomic DNA (gDNA). RNA was reverse-transcribed to cDNA using Superscript II (Thermo-Fisher Scientific, MA, USA) and gene specific primers for L1 and housekeeper TATA-binding protein (TBP) as previously described (31), (supplementary file 1). The absence of gDNA was confirmed by *HBP1* PCR and gel electrophoresis (supplementary file 2). RT-PCR using SYBR Green Master Mix (Thermo Fisher Scientific) was performed using specific primers for L1-5’UTR and TBP (supplementary file 1). Subsequent expression was displayed as a ratio of a biological control (HEK293T cell line ERE expression) to minimise any batch effects.

Sorted cell subsets were processed as previously described (29, 30). In brief, the contemporaneous lymphocyte subsets had RNA isolated using Qiagen RNeasey Plus Micro Kits which was then applied to a gDNA Eliminator spin column (both Qiagen, Germany) as per manufacturer’s instructions. For the B cell subsets, 15000 cells were sorted into RF10 (RPMI 1640 culture medium containing 10% FCS; both Sigma-Aldrich). After sorting, the cells were pelleted and lysed in Buffer RLT (Qiagen). Either 50 ng of RNA or the lysate from 15000 cells respectively was loaded onto a NanoString nCounter Human immunology V2 Panel chip (NanoString Technologies Inc., WA, USA) including customized probes against ALUYa5, AluYb9, LTR5, L1-5’UTR and L1-ORF2 (supplementary file 3) and analyzed according to manufacturer’s instructions.

### Synovial tissue

Synovial biopsy specimens of wrist/knee joints were retrieved as described (32) using a 16-gauge Quick-Core Biopsy Needle [Cook Medical] or Temno Biopsy Needle [Carefusion/Becton Dickinson]) from consenting individuals prior to the commencement of immunomodulatory therapy, including systemic glucocorticoids. Tissue was paraffin-embedded as previously described, approximately 24 hours after collection, into 10% neutral buffered formalin (33). Total RNA was extracted from curls taken from FFPE blocks using the RNeasy FFPE kit, and quality-assessed by Qubit fluorometric quantitation according to manufacturers’ instructions. Samples that passed QC (25ng) for transcriptional profiling employed the nCounter® PanCancer Immune profiling codeset panel, modified to include probes for EREs (supplementary file 3); this was carried out using the NanoString nCounter® FLEX analysis system according to the manufacturer’s instructions.

#### In silico analysis of established RA synovial tissue single cell sequencing datasets

*RepEnrich* is a computational method that allows analysis of repetitive elements in any organism with a reference genome available that has repetitive element annotation (34). This platform was applied to freely available established RA and OA synovial tissue single cell data (https://immunogenomics.io/ampra) from the Accelerating Medicines Partnership Rheumatoid Arthritis and Systemic Lupus Erythematosus Consortium (35). Full analysis details in supplementary file 4. Comparison between synovial cell subsets was performed using the cellular clustering described in (35).

### Statistical analysis

GraphPad Prism (V.5.0; GraphPad Software, La Jolla, Calif) and R Core Team (2020) software was used. Univariate generalized linear models, Mann–Whitney *U* tests, one-way ANOVA (with Tukey’s *post hoc* analysis) and Wilcoxon-signed rank tests were performed employing a significance threshold where α = 5%. Lymphocyte and B cell subsets NanoString nCounter data analysis was performed in R (v4.2.1) as described previously (29, 30). Synovium data was processed similarly, as outlined in supplementary file 5.

## Results

### Patient cohorts

The whole blood LINE1 analysis cohort included 56 patients with early RA (eRA). Simultaneous B cell, T cell, DC and monocyte cell specific retroelement expression was obtained from 8 rheumatoid factor positive and anti-CCP positive (double seropositive) eRA patients. B cell subset expression was assessed amongst double seropositive eRA and early PsA patients (n=4 each) matched for age and sex, with comparable levels of inflammation (CRP, ESR). The synovial tissue cohort comprised 22 eRA patients. Full demographical and clinical data are shown for all the cohorts in Table 1.

**Table 1:**
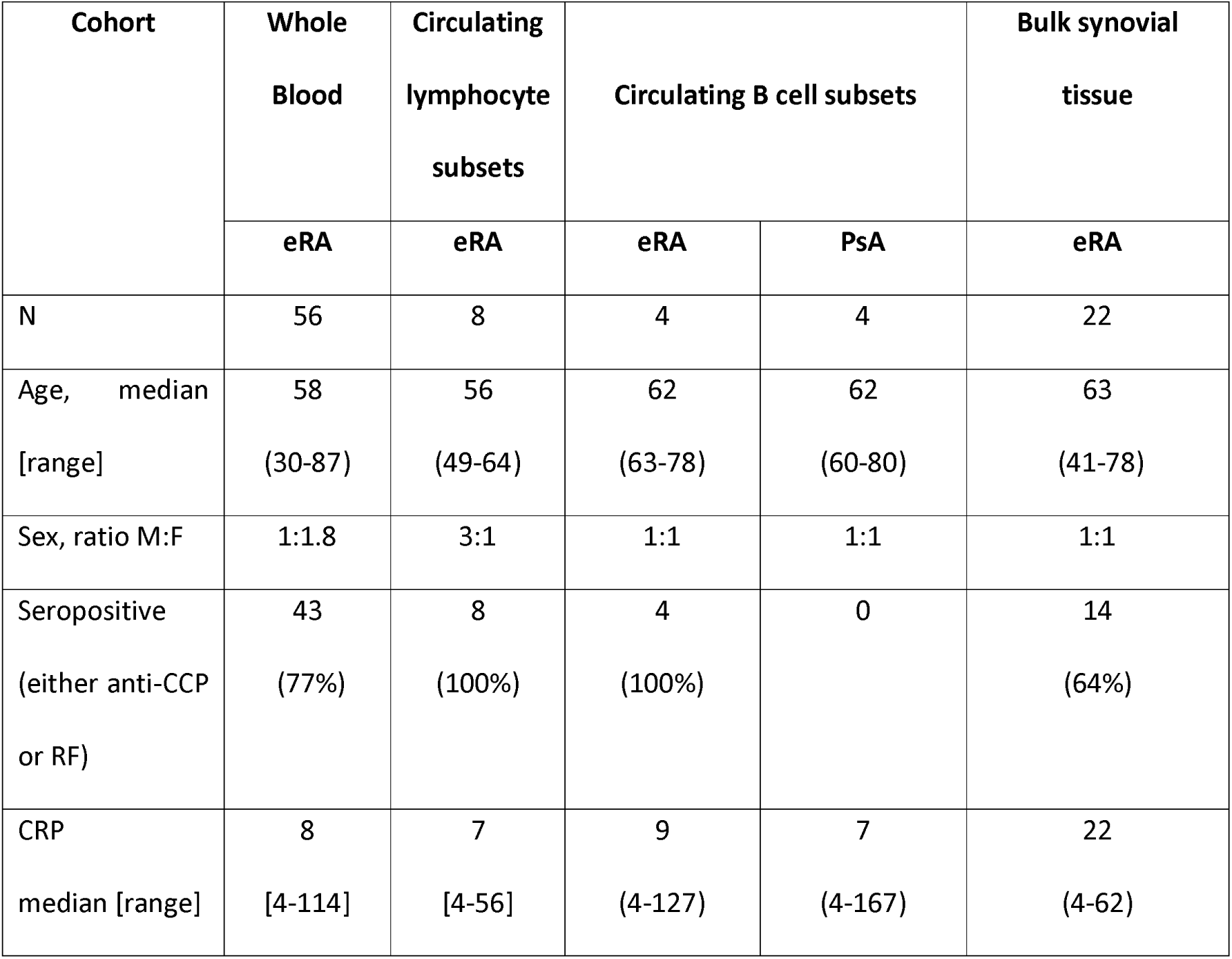
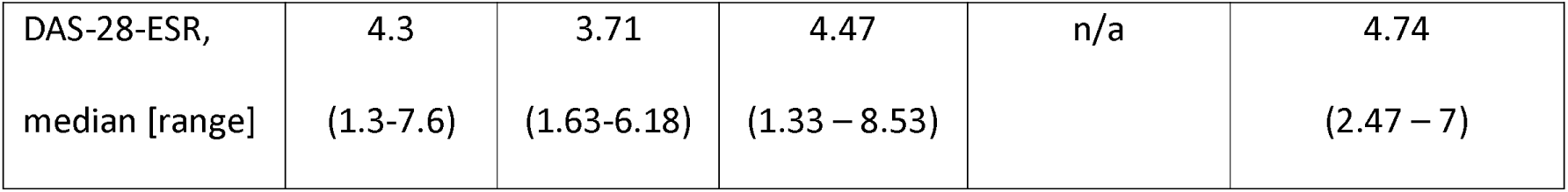
Patient and controls demographic data. Anti CCP, anti-citrullinated protein/peptide antibody; CRP, C reactive protein; DAS-28, disease activity score; eRA, early rheumatoid arthritis; ESR, erythrocyte sedimentation rate; F, female; M, male; n/a, not applicable; RF, rheumatoid factor; PsA, psoriatic arthritis

### Early RA synovial and peripheral blood endogenous retroelement expression associates with increased IFN-α

In early RA whole synovial tissue, hierarchical clustering of co-expression correlations of all available genes demonstrated clustering of *IFNA* and retroelements, figure 1A. A heatmap of the correlations between genes within the ERE cluster is shown in supplementary file 6. Pathway analysis (KEGG) of the ERE cluster implicated enrichment of JAK-STAT signalling (p=0.004), primarily due to association with IFN-I, an enrichment also seen in GO terms (p=2.71 x10^-6^), supplementary file 6. *IFNA* transcripts (*IFNA1, IFNA2, IFNA7, IFNA8* and *IFNA17)* significantly positively associated with all classes of ERE (figure 1B) but was strongest for LTR5: IFNA17, R=0.91, Pearson correlation coefficient, BH corrected p = 4.68 x10^-9^. There was no significant association with any other cytokine transcript, including IFN-γ, IL-6, IL-12 p70, TNF-α, IL-1β, IL-2, IL-13, IL-4 and IL-10 (data not shown). In early RA whole blood, there was a highly significant positive association between LINE1 transcript expression and circulating IFN-α protein, p=0.018 (figure 1C). This was not seen with any of the other circulating cytokines measured: IFN-γ, IL-6, IL-12 p70, TNF-α, IL-1β, IL-2, IL-13, IL-4 and IL-10, p>0.05 for all, data not shown. There was no significant association between the whole blood IGS and LINE1 transcript expression (figure 1D) despite a positive trend (p=0.06) between the IGS and circulating IFN-α (data not shown). Finally, whole blood LINE1 expression did not correlate with age nor sex, supplementary file 7.

**Figure 1.**
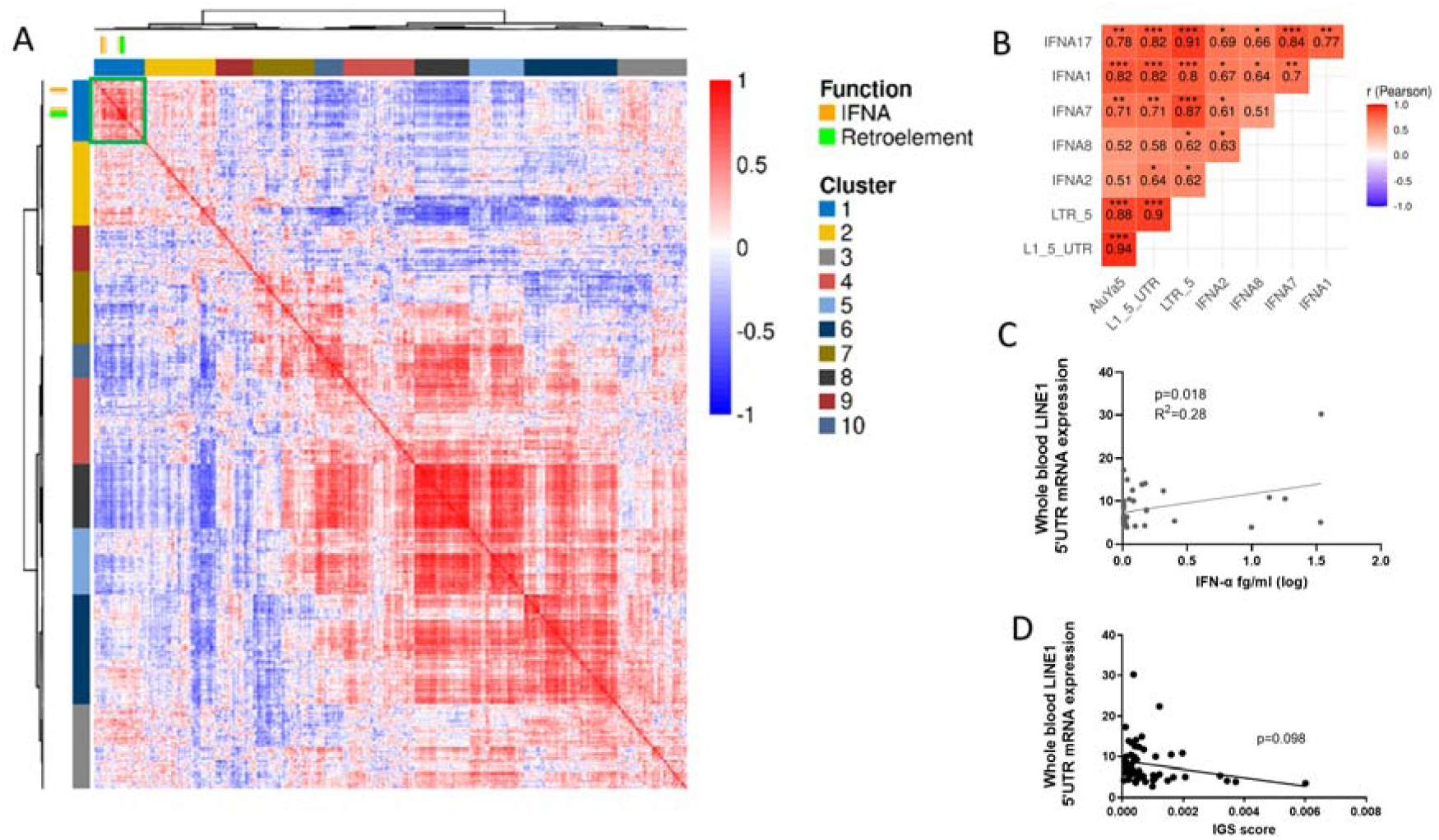
**(A)** Hierarchical clustering of early RA bulk synovial tissue gene expression correlations. Rows and columns depict genes, and the colour bar represents the Pearson’s coefficient (r) of their pairwise gene expression correlation. Dendrograms show hierarchical clustering of the genes by their expression correlation patterns. IFNA and EREs are highlighted by individual gene markers and the locations of their correlations by the green box. **(B)** Heatmap of correlation profiles between early RA bulk synovial IFN transcript and ERE classes. Significant (adjusted p value <0.05) correlations are highlighted, *p<0.05, ** p<0.01, ***p<0.001. **(C)** Whole blood LINE-1 5’UTR (L1–5’UTR) expression was analysed in early RA patients and shown in arbitrary units in relation to expression in HEK293T. Linear regression of early RA whole blood (L1-5’UTR) and circulating IFN-α protein level, n= 42 and **(D)** whole blood IGS, n=56 is shown.

### Early RA whole blood LINE1 expression correlates with anti-CCP titres

There was a significant positive association between anti-CCP titres (IU) and LINE1 (L1-5’UTR, linear regression, p<0.0001, R^2^=0.38) which was not seen for RF titres (figure 2A, 2B). Expression did not appear to reflect global B cell function as there was no association between circulating immunoglobulin levels, IgM, IgG or IGA and whole blood LINE1 (linear regression, p>0.4 for all) (figure 2C). Smoking is implicated in both anti-CCP generation and ERE activity (36, 37), however, there was no significant difference in whole blood LINE1 expression between cohorts based on smoking status (p>0.05, ANOVA), figure 2D. There was also no significant association between LINE1 expression and disease activity (DAS-28), nor its components including, TJC, SJC, VAS, CRP and ESR (data not shown).

**Figure 2.**
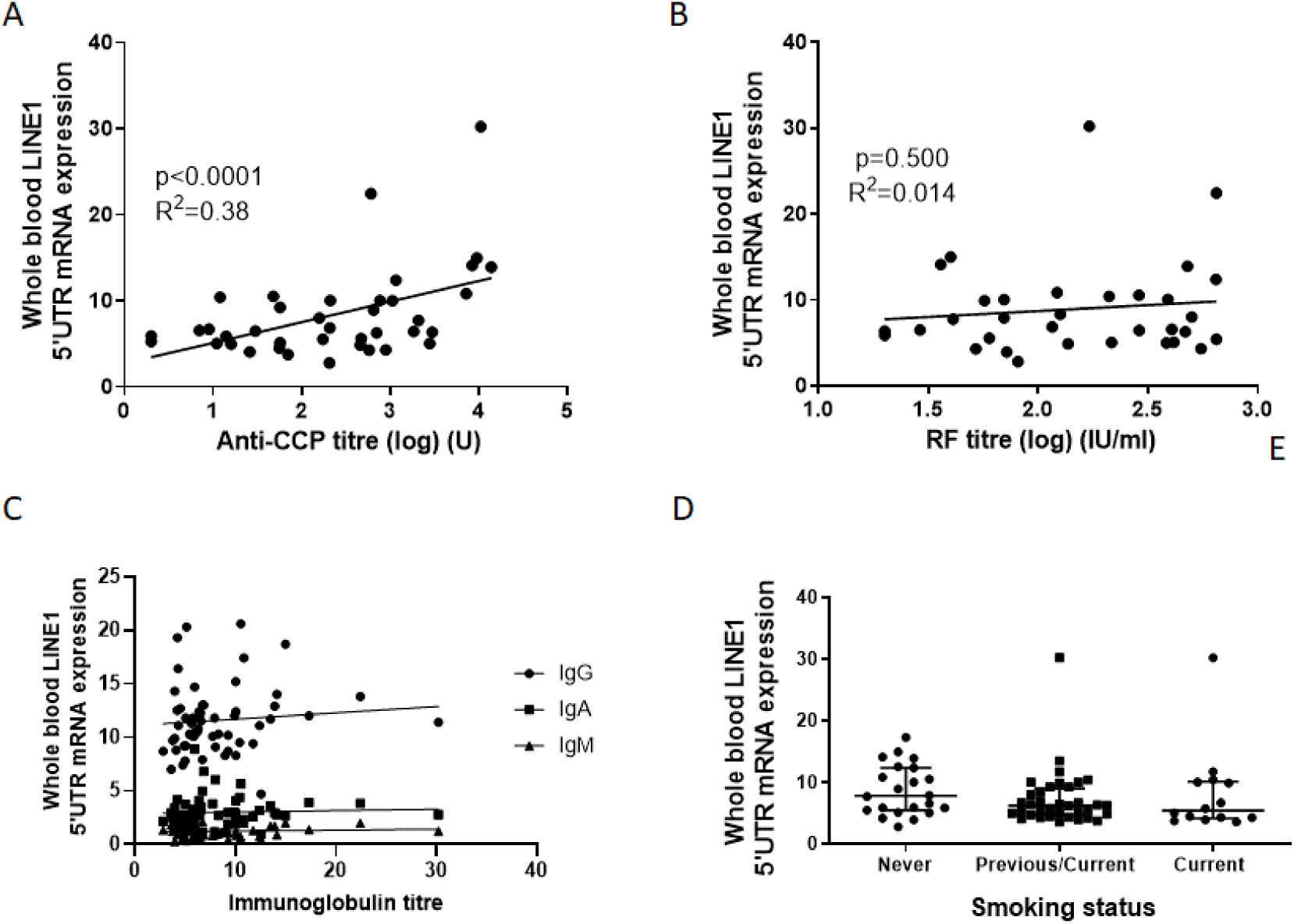
Whole blood LINE-1 5’UTR (L1–5’UTR) expression was analysed in early RA patients (n=56) and is shown in arbitrary units in relation to expression in HEK293T cells. **(A-B)** Linear regression between early RA patient (n=39) anti-CCP titres or RF titres (n =34) and whole blood L1-5’UTR expression **(C)** Linear regression between L1-5’UTR expression and circulating immunoglobulins (IgA, IgM and IgG) in early RA patients (n=56), p>0.05 for all. **(D)** Comparison of L1-5’UTR expression between early RA patients based on smoking status, never, previous/current and current.

### Circulating early RA B cells, particularly naïve and CD5 subsets, have increased ERE expression which associates with increased IFNA transcript

LTR5, LINE1 and AluYa5 expression was compared between lymphocyte subsets (B cells, pDCs, CD1c^+^ DCs, CD14^+^ monocytes, CD8^+^ and CD4^+^ T cells) from 8 double seropositive early RA (eRA) patients. ERE expression was significantly increased in B cells compared with other lymphocyte subsets (figure 3A). ERE activity in the B cell compartment was examined further comparing ABCs, naïve, memory and CD5+ B cells in eRA, with early psoriatic arthritis patients (PsA) as disease controls. There was a trend towards increased expression of ERE in eRA patients in all subsets, which became highly significant for naïve B-cells (figure 3B).

**Figure 3.**
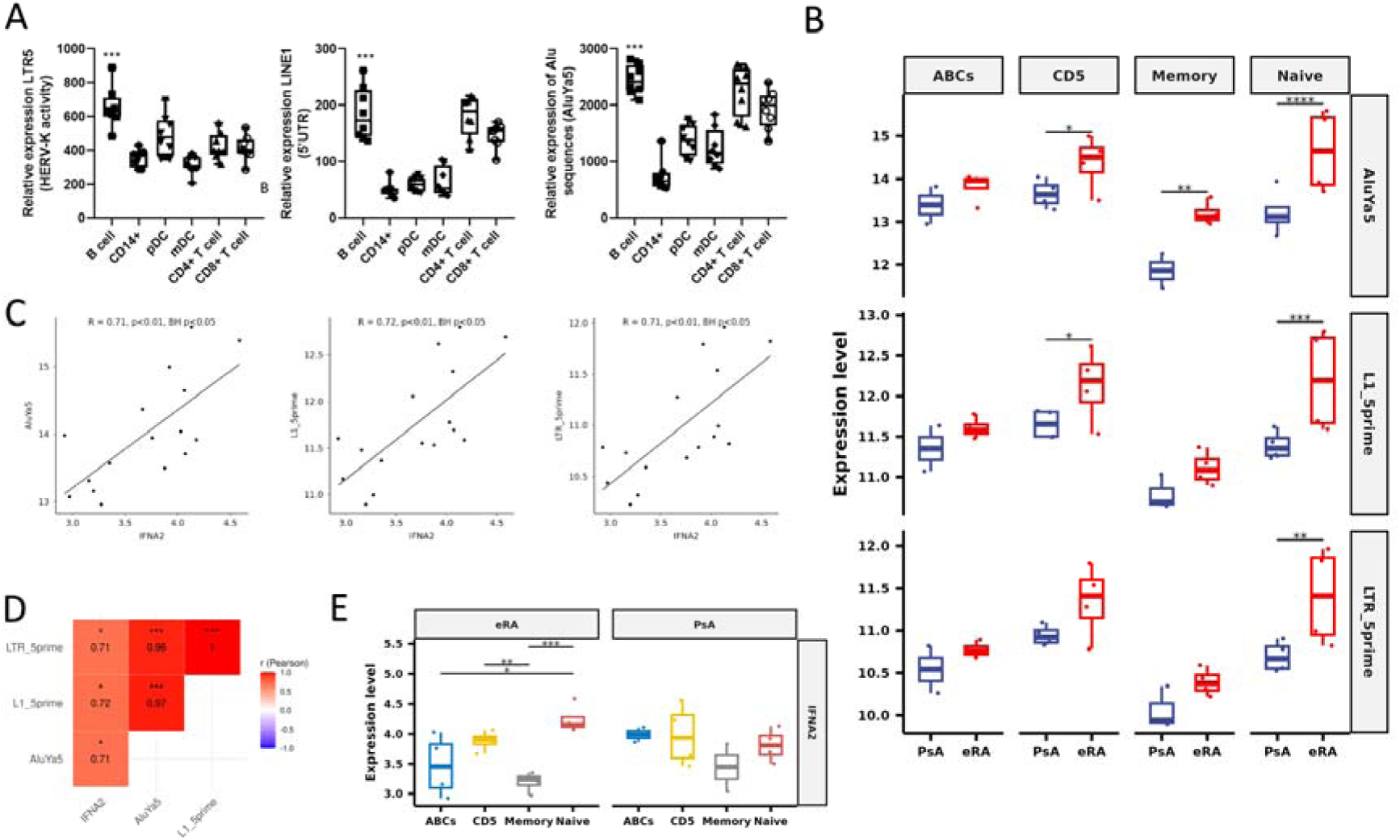
**(A)** Circulating lymphocyte (CD19^+^ B cells, CD14^+^ monocytes [CD14], plasmacytoid dendritic cells [pDCs], conventional CD1c^+^ DCs [mDCs], CD4^+^ T cells [CD4] and CD8^+^ T cells [CD8]) retroelement (ERE) expression (LTR5, LINE1 [L1-5’UTR], AluYa5) in early RA (eRA, n=8). Data are presented as box and whisker plots, in which the horizontal line represent the median value, the box represents upper and lower quartiles, and the whiskers represent range. Kruskal-Wallis test. (B) Expression of all retroelement classes in B cell subsets, age associated B-cells (ABCs, CD19^+^CD11c^+^CD21^−^), CD5^+^ B cells (CD19^+^CD5^+^), memory B cells (CD19^+^IgD^−^CD27^+^) and naïve B cells (CD19^+^IgD^+^CD27^−^) from early RA patients (eRA) and early disease controls (early psoriatic arthritis, PsA), Wald test. (C) Pearson correlation coefficient of *IFNA2* and SINE (AluYa5), LINE1 (L1_5prime) and LTR5 (LTR_5prime) expression from eRA B cells, p<0.01, BH corrected (D) Heatmap of correlation profiles between *IFN* transcript and retroelement classes in pooled eRA B cell subsets. Significant (adjusted p value <0.05) correlations are highlighted (E) Comparison of *IFNA2* expression across B cell subsets in eRA and early psoriatic arthritis (PsA), paired T tests. *p<0.05, ** p<0.01, ***p<0.001.

In eRA pooled B cells, ERE significantly associated with *IFNA* transcript (figure 3C and 3D). Hierarchical clustering of co-expression correlations of all available genes further demonstrated clustering of *IFNA* and EREs, supplementary file 8. Clusters were visually defined and heatmap of the correlations between the genes within the ERE cluster is shown in supplementary file 9. Pathway analysis of this cluster in eRA alone demonstrated limited terms achieving significance, however when examining pooled PsA and eRA data, increased ERE expression was associated with enrichment of KEGG pathways relating to viral infection as well as PI3K-Akt signalling, and GO terms were enriched for lymphocyte activation involved in immune response (p=0.0005) supplementary file 9.

Furthermore, in eRA there was significantly increased *IFNA2 t*ranscript in naïve and CD5 B cells compared with memory B cells, a pattern not seen in PsA (Figure 3F), and, when comparing directly between eRA and PsA, there was a trend towards higher expression of *IFNA2* in eRA Naïve B cells than in PsA Naïve B cells, although this was not significant (supplementary file 10). Finally, to explore potential signalling pathways we examined associations between EREs and key innate immune sensors, RIG-1, MDA5, TLR7, cGAS and TLR9 in the pooled lymphocyte subsets. A significant positive association was only seen between RIG-1 and LTR5 (R^2^ = 0.64, p<0.05), supplementary file 11.

### SAMHD1 implicated in early RA peripheral blood B cell retroelement replication

To evaluate potential mechanisms driving increased ERE transcripts in circulating B cells, we examined expression of key enzymes involved in their activation. SAM and HD Domain Containing Deoxynucleoside Triphosphate Triphosphohydrolase 1 transcript (*SAMHD1*), an enzyme limiting retroelement replication (38), was significantly reduced in early RA B cells when compared with all other circulating lymphocytes (p<0.001) (Figure 4A). Furthermore, early RA expression of SAMHD1 inversely correlated with pooled lymphocyte ERE transcript expression (Figure 4B, supplementary file 12). SAMHD1 expression was significantly reduced (p<0.01) in eRA naïve B cells compared to ABCs and a trend noted for reduced expression compared with memory and CD5+ B cells. This pattern was not seen in PsA controls (Figure 4C). Finally, SAMDH1 was uniquely and significantly reduced in early RA naïve B cells (p<0.005) compared with PsA naïve B cells (Figure 4D). Ribonuclease H degrades RNA in RNA/DNA hybrids and expression of one of its key components, Ribonuclease H2 subunit A (RNASEH2A), was similar across all lymphocyte subsets and did not correlate with ERE expression. Conversely, three prime repair exonuclease 1 (TREX1), another key enzyme negatively regulating ERE expression, varied by cell subset with lowest expression in T cells. There was an inverse association between TREX1 and ERE expression in early RA pooled lymphocytes, but expression in B cell subsets between early RA and PsA cohorts was comparable (all supplementary file 13).

**Figure 4.**
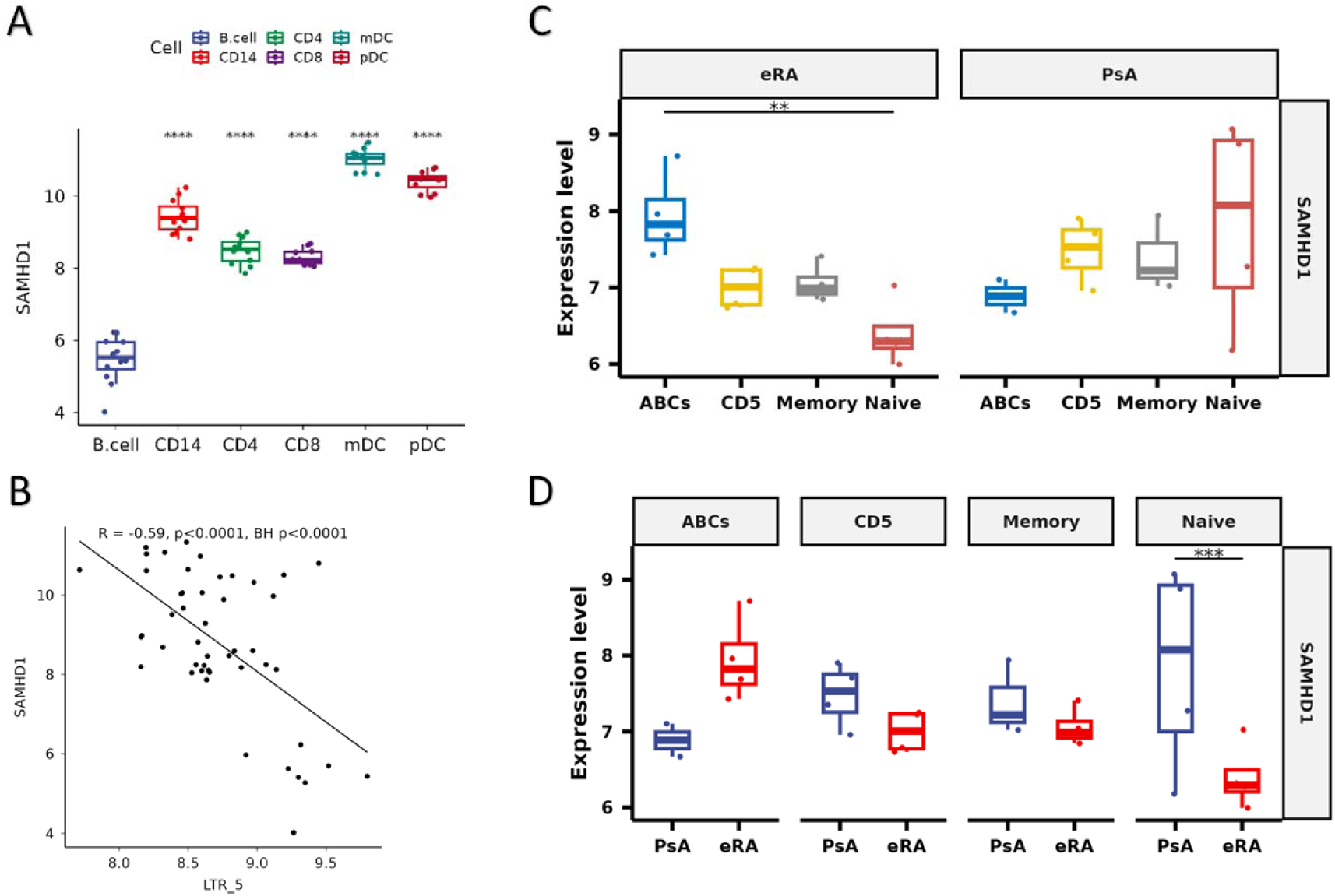
**(A)** Expression of *SAMHD1* as determined by Nanostring Technology in peripheral blood lymphocyte subsets (CD19^+^ B cells, CD14^+^ monocytes [CD14], CD4^+^ T cells [CD4], CD8^+^ T cells [CD8], conventional CD1c^+^ DCs [mDCs] and plasmacytoid dendritic cells [pDCs]) from early RA patients (n=8), B cells used as a reference in pairwise paired t tests. **(B)** Pearson’s correlation coefficient of *SAMHD1* and LTR5 in these same circulating lymphocytes pooled, BH corrected p<0.0001. **(C)** *SAMHD1* expression, determined by Nanostring Technologies, was examined in age associated B-cells (ABCs, CD19^+^CD11c^+^CD21^−^), CD5^+^ B cells (CD19^+^CD5^+^) memory B cells (CD19^+^IgD^−^CD27^+^), and naïve B cells (CD19^+^IgD^+^CD27^−^) from early RA patients (rRA) and early psoriatic arthritis patients (PsA). Differences in *SAMHD1* expression was examined within disease cohort and **(D)** between disease cohorts. Wald test with adjusted and raw P values shown for 3C and 3D respectively. ***p<0.001

Early RA bulk synovial expression of ERE did not associate with *SAMHD1, TREX1* no*r RNAseH2* and, in neither circulating early RA lymphocyte subsets, nor in synovial tissue did *DNMT1, DNMT3A* or *DNMT3B* (DNA methyltransferase enzymes, important in the epigenetic regulation of EREs) associate with ERE expression.

### The LTR class is universally increased in all RA synovial cell subsets and cluster with increased IFN-I signalling

Given differences in ERE expression observed in peripheral blood subsets, and correlation profiles between early RA bulk synovial *IFN* transcript and ERE classes (Fig 1), we wished to examine synovial tissue in more detail. Synovial single cell transcriptomic data from established RA and osteoarthritis (OA) patients was grouped into B cells, fibroblasts, monocytes and T cells as previously described (35) and reinterrogated for repeat element expression. This generated multiple individual ERE expression counts that could be grouped into class, such as LTR. Examination of the individual ERE expression counts, when grouped into class, demonstrated that in all RA synovial cell subsets, at a single cell level, LTR was proportionally the most highly expressed class (Figure 5A). Individual ERE expression counts were compared between OA and RA and the proportion of all individual ERE expression counts within each class that were comparatively either reduced or increased in RA for each cell subset demonstrated (Figure 5B and supplementary file 14). In RA, the majority of ERE expression counts within the LTR class were increased for all synovial cellular subsets, whereas counts in the LINE and SINE class were predominantly decreased. Individual ERE fold changes in supplementary data S1.

**Figure 5.**
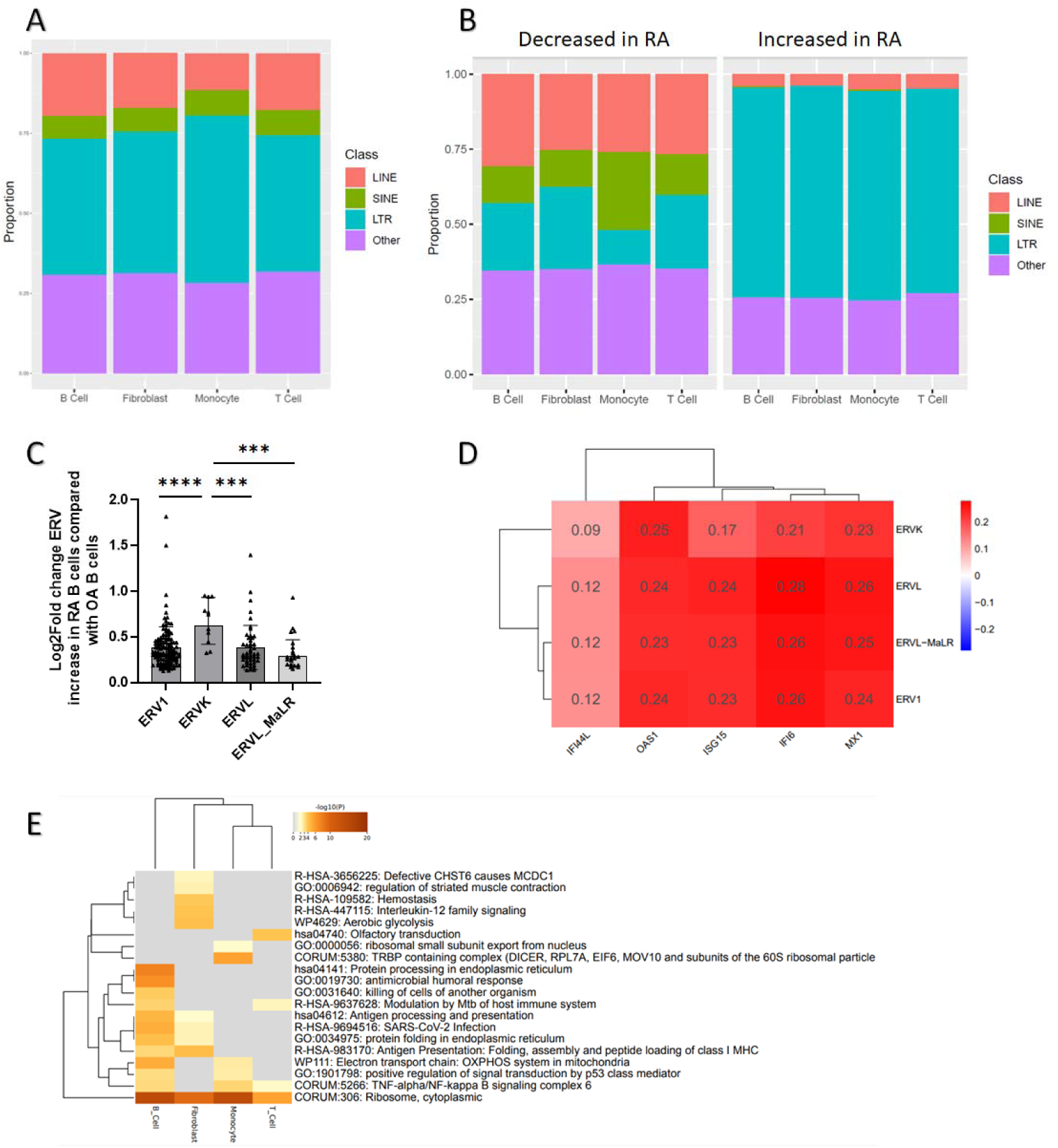
RepEnrich was applied to synovial single cell RNA-sequencing (scseq) data from established RA and non-RA controls (OA) and repeat element enrichment/endogenous retroelements (ERE) were identified. Cellular clusters included monocytes, fibroblasts, T cells and B cells. **(A)** The proportion of scseq repeat element/ERE expression, grouped by ERE class, in each cell subset in established RA synovial tissue **(B)** Proportion of individual EREs, grouped by class, with either increased or reduced expression in RA versus OA **(C)** The LTR class is divided into ERV1, ERVK, ERV, ERV_MaLR families, with individual ERE expression counts within each family. Depiction of increased differential expression (Log2FoldChange) in RA compared with OA of each individual ERE count within each ERV family. The relative increased expression (Log2FoldChange) in RA is further compared between the individual ERV families, Kruskall-Wallis (p<0.0001) and Mann-Whitney U tests **(D)** Hierarchical clustering of correlations between gene expression of interferon response genes and repeat element enrichment counts in RA B cells. Pearson’s correlation co-efficients depicted. All correlations were significant, p<0.001. **(E)** MetaScape pathway analysis of all genes with a correlation of ≥0.4 with LTR repeat elements in each of the individual cell subsets. Top 20 pathways depicted. **** p<0.0001 *** p<0.005

Given that the LTRs were most widely increased in RA we examined this class in more detail. LTRs consist of endogenous retroviruses (ERVs) families, ERV1, ERVK, ERVL, ERVL-MaLR, and individual ERE counts within these families were compared between RA and OA. Individual ERE expression counts increased in RA B cells compared with OA for each ERV family is shown in Figure C. When comparing the comparative increased counts (Log2Fold) in RA versus OA between the ERV families, overall expression was greatest in the ERVK family (Kruskal Wallis, p<0.0001), Figure 5C.

For all ERV families, hierarchical clustering of correlations between gene expression and ERE expression counts was performed. In all cell subsets there was a positive correlation between IFN-I response genes (*IFI44L, OAS1, IFI6, ISG15* and *Mx1)* and LTR expression grouped by ERV family. All correlations met statistical significance with p<0.001. Figure 5D depicts B cell correlations (supplementary file 15 for remaining cell subsets). *IFNA* counts were too low for comparable analysis to be performed. Pathway analysis of the top 20 pathways corelating with LTR repeats across cell subsets demonstrated enrichment of viral response (SARS-CoV-2), antigen processing pathways and antimicrobial humoral response in B cells (Figure 5E)

## Discussion

We examined endogenous retroelements (EREs) for the first time in drug naïve early RA and demonstrate that EREs are transcriptionally active in both whole blood and synovial tissue, with variable expression across circulating lymphocyte subsets. Expression was highest in B cells, particularly naïve B cells, which was not seen in PsA. We demonstrate for the first time that ERE activity in early RA blood and synovial tissue associates with increased IFN-α at both transcript and protein level. We also saw a marked positive association between ERE and CCP titres. Cumulatively, these data suggest that EREs may contribute to RA pathophysiology.

In eRA bulk synovial tissue we identified a significant positive association between *IFNA* transcript and ERE expression, particularly noted with LTR5. LTR5 expression denotes ERVK (HML-2) activity (39) which has been recently integrated into the human genome, and multiple copies possess potential biological activity (40). Indeed, some of the most compelling evidence of the involvement of EREs in RA has implicated this ERV, which has been detected in RA patient plasma, higher levels associating with active disease (41). Although ERVK transcripts have previously been detected in established RA blood and synovial tissue (41, 42), this is the first time they have been demonstrated in early disease and in association with IFN-α upregulation. Furthermore, when re-analysing an independent, established RA synovial tissue scSeq dataset in the public domain, we observed ERVK to be the most upregulated subtype when compared with other ERVs. In contrast however to early RA bulk synovial tissue analyses, we could find no direct association between ERE expression and *IFNA* transcript in scSeq data from major cellular subsets in established RA, although there was evidence of increased downstream IFN-I signalling. We previously showed circulating IFN-α declines during the first 6 months after RA diagnosis (7), potentially explaining this difference from early RA, although the influence of distinct cellular composition and/or sampling technique cannot be excluded. ERVs also induce TNF-α (43), which induces IRG expression independently of IFN-I in RA synovial fibroblasts, an association we also identified in our pathway analysis (44) and this process may become dominant in established disease. Nonetheless, the clear association of ERE expression in RA synovial tissue with both *IFNA* transcript and IFN-I signalling mirrors what is seen elsewhere in autoimmunity (21) and warrants further exploration.

Although examining RA synovial tissue is optimal, circulating immune cell subsets may contribute to, or reflect, tissue specific processes. Notably, early RA synovial fluid IFN-α levels are comparable to those in the circulation (7). In early RA, we demonstrated positive associations between EREs and circulating B cell *IFNA* transcript levels as well as between whole blood *LINE1* activity and circulating IFN-α protein levels. The dsRNA sensor RIG-1 was potentially implicated in ERE sensing by these data, consistent with our previously reported association between RIG-1 and circulating IFN-α levels in early RA (7). An association between *IFNA* transcript and whole blood retrotransposon activity has recently been reported in other autoimmune diseases (45) but our demonstration of an association with IFN-α protein reinforces potential biological relevance in RA. Similar to other autoimmune diseases (45), we did not find a significant association between retrotransposon activity and downstream IFN-I signalling, although we noted a trend towards an inverse association with the IGS. This may be because many IRGs are potent LINE1 negative regulators (46) and thus examining protein levels, where possible, may be optimal.

This is the first time all major classes of EREs have been simultaneously examined in circulating lymphocyte subsets, wherein we found highest ERE expression in B cells, particularly naïve B cells. Although background inflammation levels could affect ERE activity (27), expression levels were increased in RA B cell subsets compared with early PsA patients matched for inflammation. Furthermore, ERE expression in eRA whole blood was independent of other circulating inflammatory cytokines. In keeping with this differential ERE expression, there was increased *IFNA* transcript in RA naïve B cells. Single cell analysis of RA circulating B cell subsets previously demonstrated increased sensitivity to IFN-α and increased *IFNA* transcript in RA naïve B cells, resulting in increased basal activation and proliferation (47). The role of IFN-α in B cell function and pathophysiology of autoimmunity has been well established whereby it can enhance B cell proliferation, activation and autoantibody production (3). Pre-treatment with IFN-α also enhances pathological B cell proliferative responses and plasmablast differentiation (48). This potentially associates EREs to known RA pathophysiological processes via enhanced IFN-I signalling. EREs may also be implicated in B cell driven autoimmunity independently of IFN-α, via antibody responses to cell components associated with ERE nucleic acid, allowing molecular mimicry and cross activation to occur (24, 49). Overlap between rheumatic disease associated autoantibodies, including anti-Ro60 and RF, have been linked to ERE activity (26, 27, 50). We also showed a significant positive association between whole blood LINE1 activity and anti-CCP titres, and our pathway analysis suggested a positive correlation between synovial B cell EREs and antigen processing and presentation. Antibodies against HERV-K *env,* as well as against its citrullinated form, have been detected in established RA, are increased in ACPA+ patients, and positively correlate with CCP titres (51). Together these data hint at a role for EREs in promoting B cell dysfunction in RA.

Variation in circulating B cell ERE expression was associated with a reciprocal decrease in *SAMHD1*. This enzyme depletes intracellular dNTP pools thus limiting ERE replication and its deficiency has been implicated in interferonopathies (52). SAMHD1 expression can vary between cell subsets (53) and, in B cells, is increased in G1 cell cycle phases where it can enhance the development of high affinity antibodies (54). Naïve B cells, in phase G0, theoretically therefore would have lower levels of *SAMHD1* and thus increased ERE expression, as we demonstrated. We also saw enrichment of P13k/AKT signalling in B cells, and in AGS this pathway has been implicated in linking SAMHD1 deficiency to increased IFN-I response (55). *SAMHD1* expression levels do not necessarily correlate with its dNTPase activity and cellular dNTP availability (53), nevertheless the reciprocal variability in expression levels between ERE expression and *SAMHD1* are suggestive that an association may exist.

We did not see any link between SAMHD1 expression and synovial ERE expression, and other retrotransposon regulatory mechanisms may be more relevant here, such as epigenetic silencing (21, 56), promoter methylation having been shown to affect LINE1 activity in autoimmune diseases (21, 45). Although we did not explore these mechanisms in detail, we did not see any difference between retrotransposon activity and DNMT1, DNMT3A or DNMT3B expression, known epigenetic modifiers of ERE activity (45). This may reflect sample size but may also suggest other mechanisms are dominant, such as SAMHD1 in B cells, or even differential expression in the recently described HUSH complex, a known gatekeeper of ERE induced IFN-I expression (56). These provide promising avenues for future research.

In conclusion, we examine for the first-time endogenous retroelement (ERE) activity in early RA and present potentially important associations between ERE activity, IFN-I, B-cell function and autoantibody generation. Within this context it is intriguing to note that antiretroviral drugs (HAART) have ameliorated symptoms in RA (57) and further work is needed to comprehensively explore the putative pathogenic involvement of EREs in RA. This will allow greater understanding of RA pathophysiology and potentially provide new targets for disease treatment or even disease prevention.

## Contributors

FAHC, RS and JDI designed the study. FAHC, GVP, BM, VB and DD participated in data and sample collection. FAHC, RS, NN, AGP, FZ, JC, CMUH and AEA assisted with data analysis and interpretation. FAHC wrote the first draft, and all authors approved the final manuscript. FAHC is guarantor of this study.

## Data Availability

The data are available for the purposes of academic research on reasonable request to the corresponding author.

## Acknowledgements

Newcastle researchers received infrastructural support via the Research into Inflammatory Arthritis Centre Versus Arthritis (RACE) (grant number 22072); the National Institute for Health and Care Research (NIHR) Newcastle Biomedical Research Centre for Ageing and LongTerm Conditions. The authors wish to thank Professors Stefan Siebert and Iain McInnes (University of Glasgow, UK) and Drs. Shaun Flint and Katherine Nevin, GSK) and Prof Soumya Raychaudhuri (Harvard Medical School, USA), for their contributions.

## Ethics statements

For the early disease data all patients provided written, informed consent to participate in the study, which was approved by the Northeast – Newcastle and North Tyneside 2 Research Ethics Committee (12/NE/0251). For established RA and OA data consent was obtained as outlined in (35).

## Key Messages

### WHAT IS ALREADY KNOWN ON THIS TOPIC

- Type 1 Interferons (IFN-I) are of increasing interest in autoimmunity and, in some diseases, endogenous retroelements have been implicated in IFN-I generation.
- What drives IFN-I production in early rheumatoid arthritis (eRA), and whether endogenous retroelements are involved, is unclear.

### WHAT THIS STUDY ADDS

- Retroelements are associated with increased IFN-I protein and transcript levels in eRA blood and synovial tissue. They are also strongly associated with ACPA titres and preferentially increased in eRA naïve B cells.

### HOW MIGHT THIS STUDY AFFECT RESEARCH, PRACTICE OR POLICY

- This work supports retroelements as a potential contributor to eRA pathophysiology and may provide a new target for innovative therapeutic strategies.

