## Supplementary files for "Endogenous retroelement activation is implicated in IFN-α production and anti-CCP autoantibody generation in early RA"

**Supplementary file 1**

**Whole blood primers sequences**

Whole blood cDNA generation included the gene specific primers that specifically reverse transcribed LINE1 elements and the housekeeper gene TATA–binding protein (TBP). Sequences shown below.

| Gene specific Primer | Sequence |
|---|---|
| **LINE-1 elements** | ACATGTGCACATTGTGCAGGTTAG |
| **TATA –binding protein (TBP)** | GGTGCAGTTGTGAGAG |

Primer sequences used for qRT-PCR quantification of LINE-1 (L1) elements including house keeper TATA-binding protein (TBP)

| Gene or L1 element | Primer | |
|---|---|---|
| | Forward | Reverse |
| ***L1-5'UTR*** | ACAGCTTTGAAGAGAGCAGTGGTT | AGTCTGCCCGTTCTCAGATCT |
| ***TBP*** | GAACATCATGGATCAGAACAACAG | ATAGGGATTCCGGGAGTCATG |

**Supplementary file 2**

**Validation of gDNA contamination elimination from whole blood LINE1 analysis**

RNA was extracted from whole blood using the Tempus RNA system. cDNA was generated and a PCR performed using primers which spanned an intron and exon region the HBPI gene.

| HBPI Primer sequence for c-DNA PCR | Sequence |
|---|---|
| **Forward Primer** | TCGAAGAGTGAACCAGCCTT |
| **Reverse Primer** | GAAGGCCAGGAATTGCACCATCC |

The PCR products were electrophoresed through a 1% weight/volume agarose gel containing ethidium bromide (10 mg/ml; Sigma) at a concentration of 1 µl/100 ml agarose and UV visualised using a G:BOX gel doc system (Syngene, Cambridge, UK). A 2-Log DNA ladder (New England Biolabs, MA, USA) was used for the whole blood c-DNA super-clean protocol products where contaminating DNA will produce 2 bands (570bp and 152bp), while uncontaminated cDNA will have just one product (152bp). A GeneRuler[TM] 100bp and 1Kb DNA ladders (Thermo Fisher Scientific) was used to assess the band sizes for the cell subset specific RNA PCR products, where a detectable band at 1011bp indicated gDNA contamination.

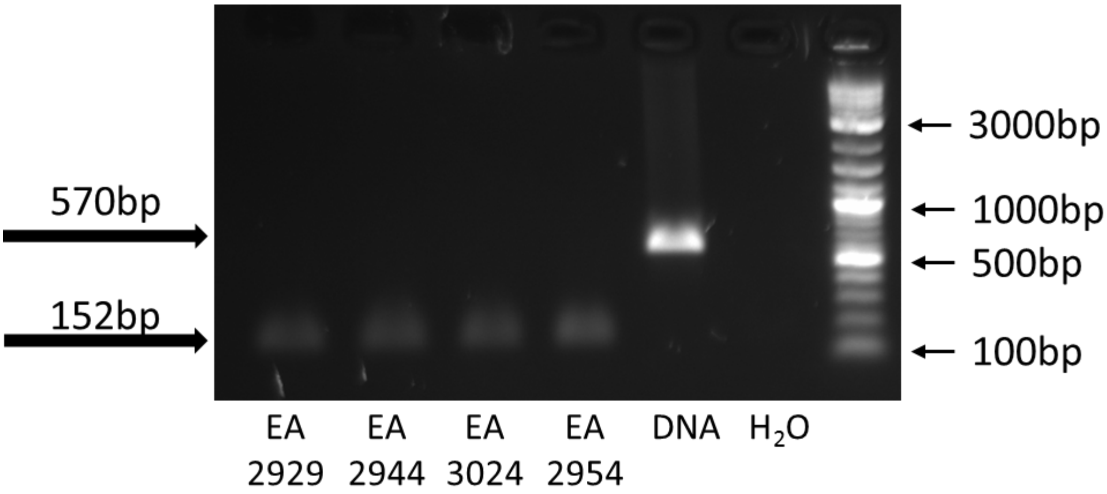


This demonstrates representative findings using patient samples (EA2929, EA2944, EA3024 and EA2954) as well as a positive DNA control and confirms successful removal of any genomic DNA.

**Supplementary file 3**

| Customised probe | Sequence |
|---|---|
| **AluYa5** | GGAAGAAGGGGGCCGGGCGCGGTGGCTCACGCCTGTAATCCCAGCACTTTGG GAGGCCGAGGCGGGCGGATCACGAGGTCAGGAGATCGAGACCATCCCGGCT AAAACGGTGAAACCCCGTCTCTACTAAAAATACAAAAAATTAGCCGGGCGTAG TGGCGGGCGCCTGTAGTCCCAGCTACTTGGGAGGCTGAGGCAGGAGAATGG CGTGAACCCGGGAGGCGGAGCTTGCAGTGAGCCGAGATCCCGCCACTGCACT CCAGCCTGGGCGACAGAGCGAGACTCCGTC |
| **L1 5'UTR** | GGGGGAGGAGCCAAGATGGCCGAATAGGAACAGCTCCGGTCTACAGCTCCCA GCGTGAGCGACGCAGAAGACGGTGATTTCTGCATTTCCATCTGAGGTACCGG GTTCATCTCACTAGGGAGTGCCAGACAGTGGGCGCAGGCCAGTGTGTGTGCG CACCGTGCGCGAGCCGAAGCAGGGCGAGGCATTGCCTCACCTGGGAAGCGC AAGGGGTCAGGGAGTTCCCTTTCCGAGTCAAAGAAAGGGGTGACGGACGCA CCTGGAAAATC |
| **LTR5 5' end** | AAGGGGGAAATGTGGGGAAAAGCAAGAGAGATCAGATTGTTACTGTGTCTGT GTAGAAAGAAGTAGACATAGGAGACTCCATTTTGTTATGTACTAAGAAAAATT CTTCTGCCTTGAGATTCTGTTAATCTATGACCTTACCCCCAACCCCGTGCTCTCT GAAACATGTGCTGTGTCCACTCAGAGTTGAATGGATTAAGGGCGGTGCAGGA TGTGCTTTGTTAAACAGATGCTTGAAGGCAGCATGCTCCTTAAGAGTCATCAC CACTCCCTAATCTCAAGTACCCAGGGACACAAAAA |

Sequences of customised probes used in NanoString panel plus nCounter to identify endogenous retroelement activity.

**Supplementary file 4**

RepEnrich2 is a method to estimate repetitive element enrichment using high-throughput sequencing data, and is a continuation of RepEnrich (Ref 1).

Briefly, the RepEnrich workflow consists of (1) align reads to the unmasked genome and divide into uniquely mapping and multi-mapping reads (2) test uniquely mapping reads for overlap with repetitive elements, while aligning multi-mapping reads separately to repetitive element assemblies representing individual repetitive element subfamilies (3) combine the counts from uniquely mapping reads and multi-mapping reads to estimate enrichment.

Alignment of fastq files to the hg38 genome was carried out using bowtie2 version 2.3.5 (Ref 2). Repetitive element annotation was obtained from the RepEnrich2 GitHub repository (https://github.com/nerettilab/RepEnrich2) and the workflow was implemented using python scripts provided by the software maintainer.

Differential analysis of repeat element enrichment analysis was carried out using the R package DESeq2 version 1.38.2 (Ref 3) using the repeat element fraction count files produced by RepEnrich2. DESeq2 estimates fold change between experimental conditions (RA samples vs non-RA samples) using a Negative Binomial GLM with a logarithmic link function.

Gene level counts were obtained by aligning fastq files to the hg38 transcriptome using Salmon version 1.9.0 (Ref 4). Gene level differential expression analysis was carried out using DESeq2.

**Supplementary file 5**

Synovium NanoString nCounter raw count data was normalized with respect to library size and subject to a variance stabilizing transformation using the vst function of the DESeq2 R package. The removeBatchEffect function of the limma R package was used to adjust for the variability due to multiple expression profiling runs. Pairwise linear associations between all genes of the NanoString panel were quantified by calculating the Pearson's correlation coefficients (r) of their expression profiles. Corresponding p values were corrected for multiple testing using the Benjamini-Hochberg (BH) method. Correlations were considered significant if adjusted p-value<0.01. The NMF package was used for the hierarchical clustering and visualization of the correlation matrix heatmaps. Euclidean distance and Ward method were applied. The clusters were defined by visual inspection of the resulting dendrograms, and their gene lists were extracted using the cutree function. Gene ontology (GO) biological process and KEGG pathway enrichment analyses of the resulting clusters were performed on the Database for Annotation, Visualization and Integrated Discovery (DAVID 2021), using all the genes of the NanoString panel as background gene list.

**Supplementary file 6**

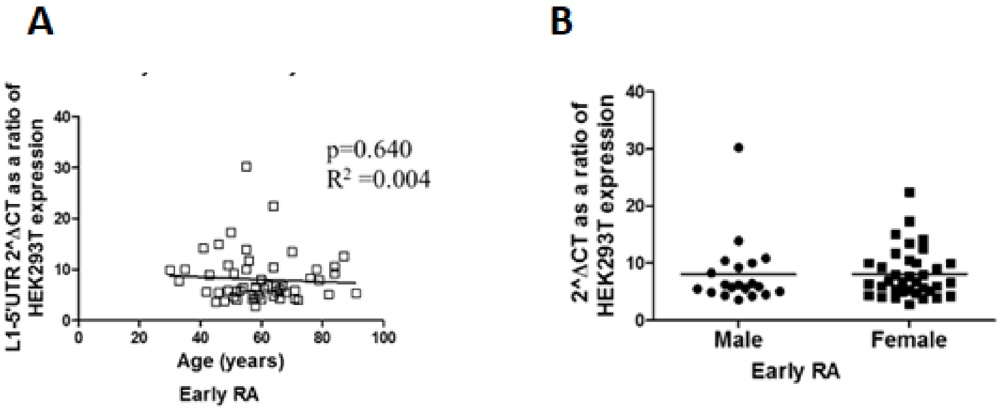


Whole blood LINE-1 expression (L1-5’UTR) was analysed in early RA patients (n=56). **(A)** Associations with age were examined. Plots depict linear regression between age and L1-5’UTR expression in early RA. **(B)** Differences in LINE1 expression with relation to gender were examined in early RA. Horizontal lines depict median values. No comparison met statistical significance (Mann-Whitney U test, where significance $p<0.05$).

**Supplementary file 7**

A

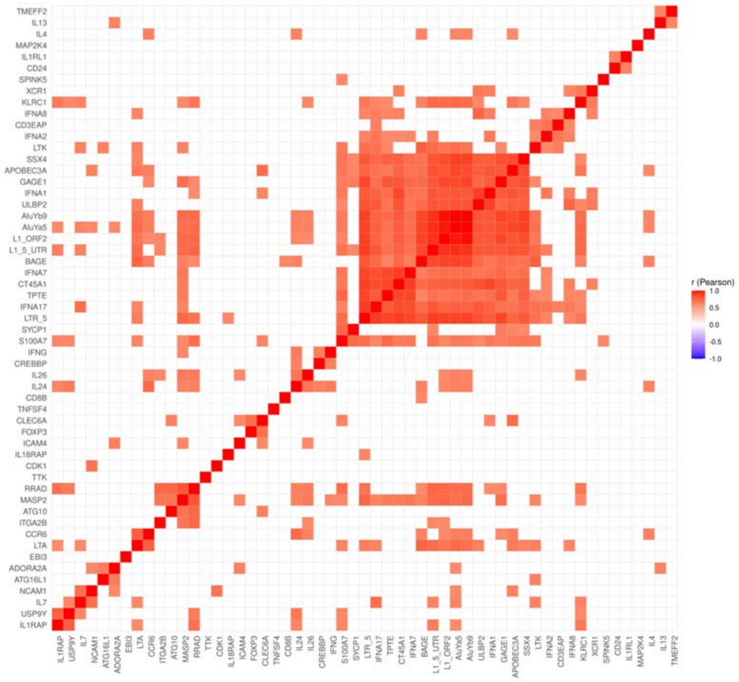

B

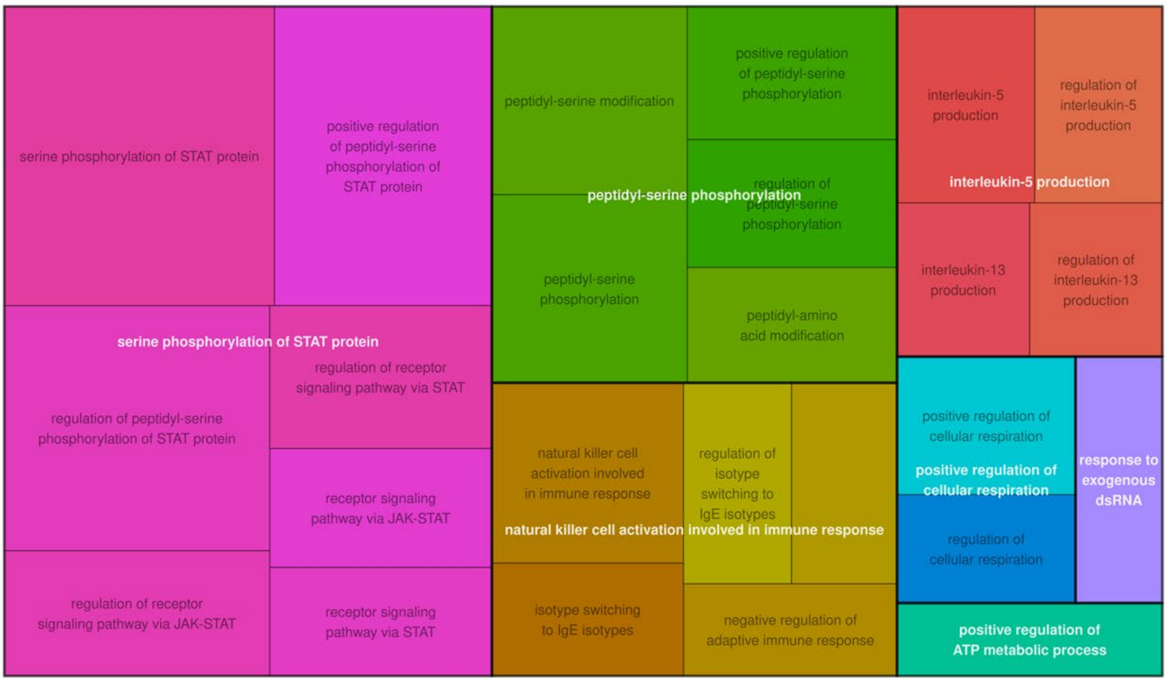


**A.** Heatmap of the pairwise gene expression correlation matrix which included genes with the most similar correlation profiles to the retroelements in bulk early RA synovial tissue, as highlighted in figure 1D. Correlations were reordered using hierarchical clustering and the insignificant ones (adjusted p>0.05) display a blank square. **B** Treemap plots showing the enriched GO terms (raw p < 0.05), within the cluster in (A) grouped based on their semantic similarity. Groups were coloured and named based on the term with the highest score (-log10(raw p)). The space used by each term is proportional to its score.

**Supplementary file 8**

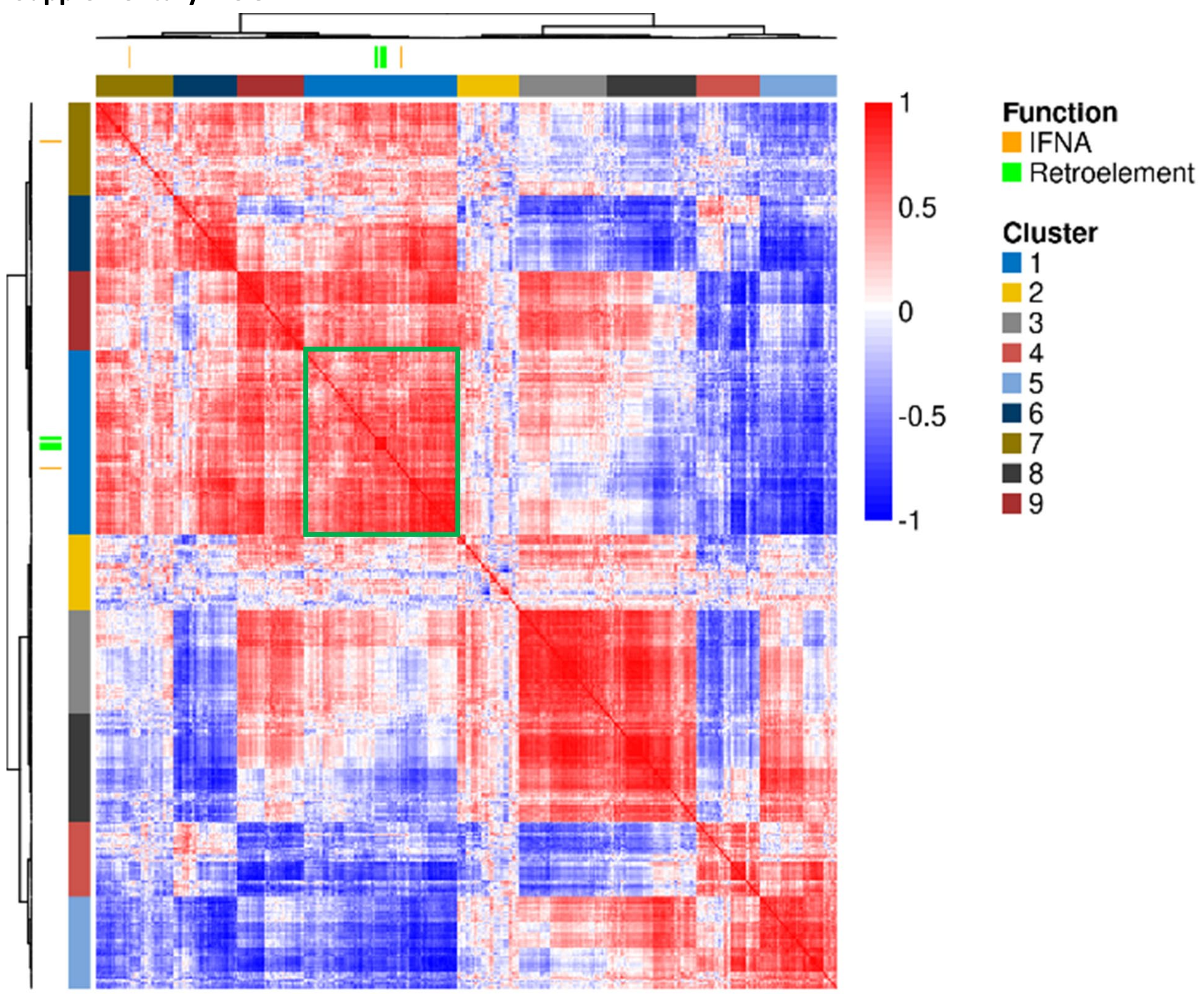


Hierarchical clustering of early RA circulating B cell gene expression correlations. Rows and columns depict genes, and the colour bar represents the Pearson's coefficient (r) of their pairwise gene expression correlation. Dendrograms show hierarchical clustering of the genes by their expression correlation patterns. IFNA and retroelements are highlighted by individual gene markers and the locations of their correlations by the green box.

### Supplementary file 9

A

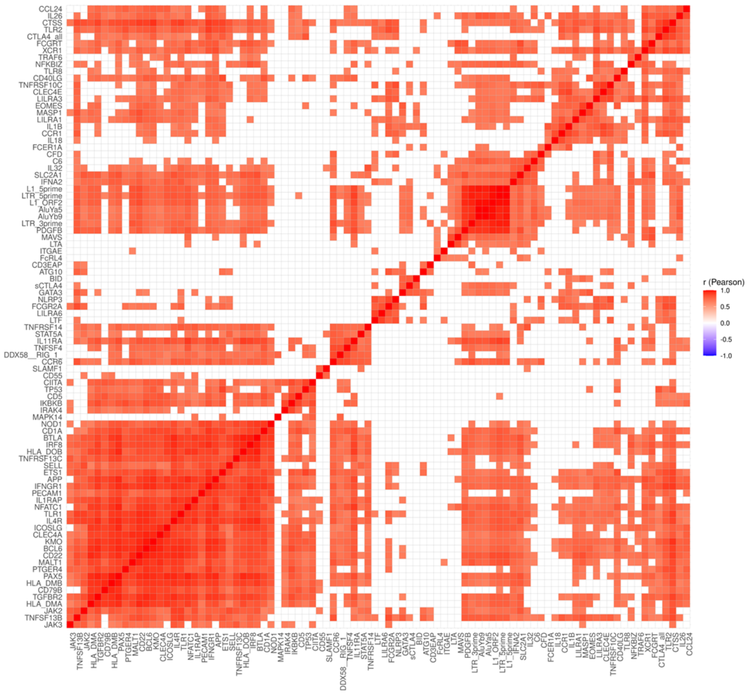


B

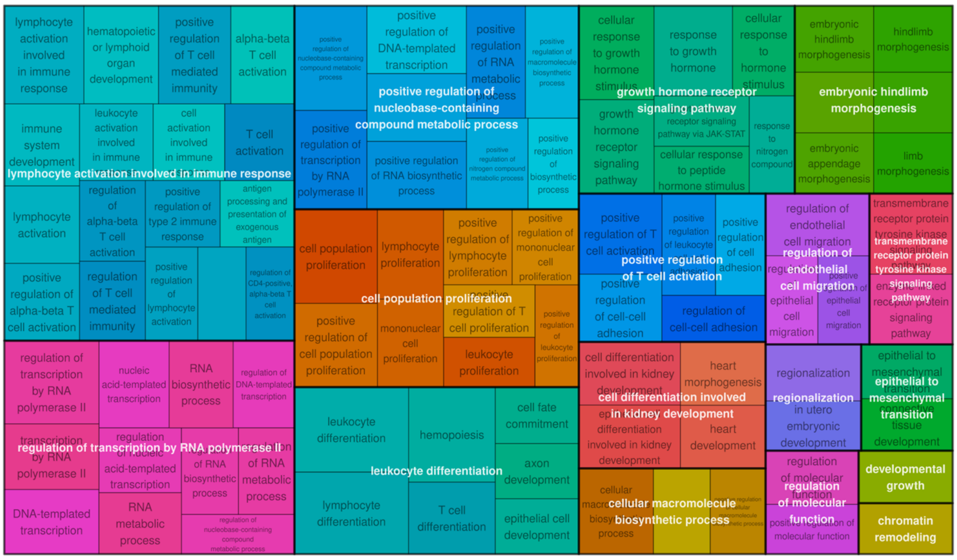


C

| KEGG term | KEGG Pathway | Gene Count | P Value |
|---|---|---|---|
| hsa05145 | Toxoplasmosis | 14 | 0.00036 |
| hsa04658 | Th1 and Th2 cell differentiation | 14 | 0.00082 |
| hsa05164 | **Influenza A*** | 16 | 0.00128 |
| hsa05169 | **Epstein-Barr virus infection*** | 16 | 0.00678 |
| hsa04151 | PI3K-Akt signalling pathway | 11 | 0.01769 |
| hsa05206 | MicroRNAs in cancer | 7 | 0.02396 |
| hsa04935 | Growth hormone synthesis, secretion and action | 5 | 0.02747 |
| hsa05200 | Pathways in cancer | 16 | 0.02926 |
| hsa05161 | **Hepatitis B*** | 12 | 0.03245 |
| hsa05416 | **Viral myocarditis*** | 7 | 0.04070 |
| hsa05168 | **Herpes simplex virus 1 infection*** | 14 | 0.04362 |
| hsa04550 | Signalling pathways regulating pluripotency of stem cells | 5 | 0.04541 |
| hsa05340 | Primary immunodeficiency | 7 | 0.04761 |

**A.** Heatmap of the gene-to-gene correlations matrix which included genes with the most similar correlation profiles to the retroelements in pooled early RA B cell subsets, as highlighted in supplementary file 8. Correlations were reordered using hierarchical clustering and the insignificant ones (adjusted $p>0.05$) display a blank square. **B** Treemap plots showing the enriched GO terms (raw $p < 0.05$), and **(C)** KEGG pathways analysis within the equivalent of the cluster in (A) for pooled early RA and PsA samples, as grouped based on their semantic similarity. For Treemap plots groups were coloured and named based on the term with the highest score (-log10(raw p)). The space used by each term is proportional to its score. In the KEGG pathway ***** depicts pathways known to associate with viral infection.

**Supplementary file 10**

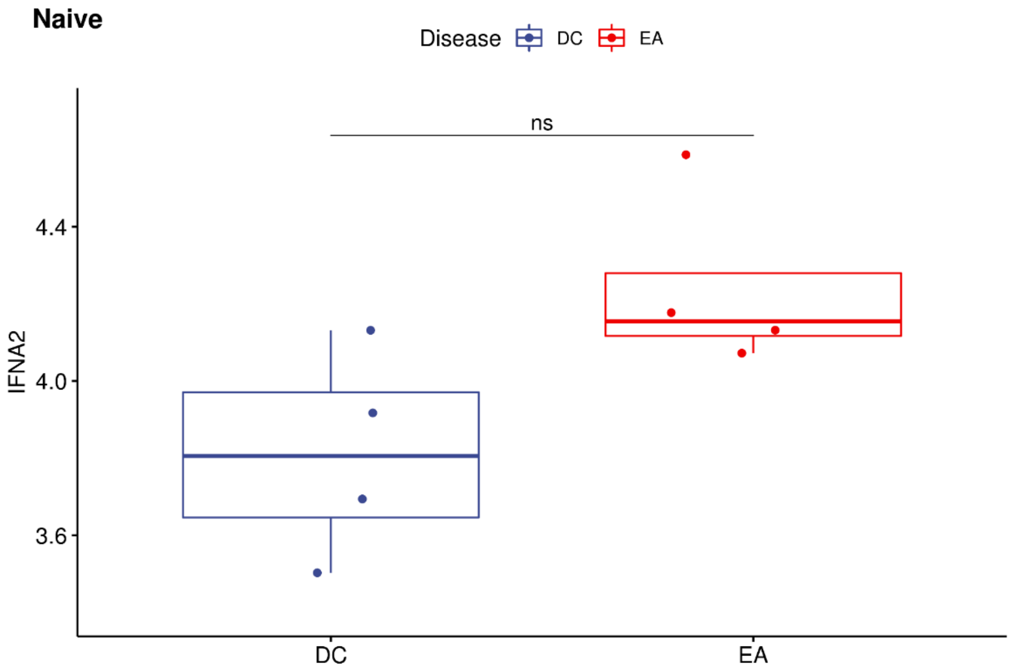


Flow cytometrically sorted naïve B cells (CD19$^+$IgD$^+$CD27$^-$), from both early RA patients (EA) and disease control, early psoriatic arthritis patients (DC), were assessed by Nanostring to determine differences in *IFNA2* expression. Wald test with adjusted P values shown.

**Supplementary file 11**

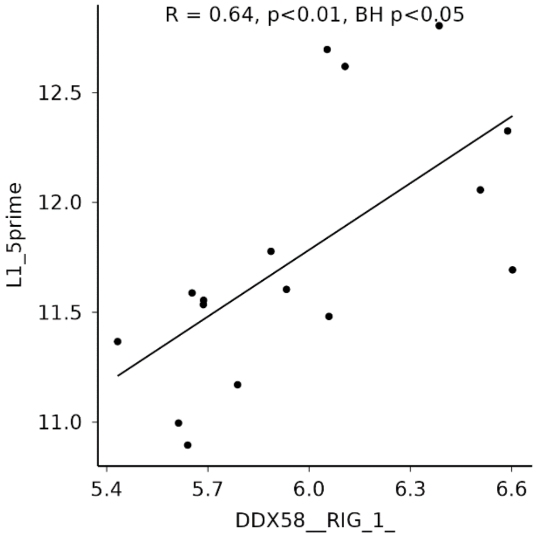


Gene expression correlation between *RIG1 (DDX58_RIG_1)* and LINE1 (L1_5_UTR) in peripheral blood pooled B cell subsets. R: Pearson's correlation coefficient.

**Supplementary file 12**

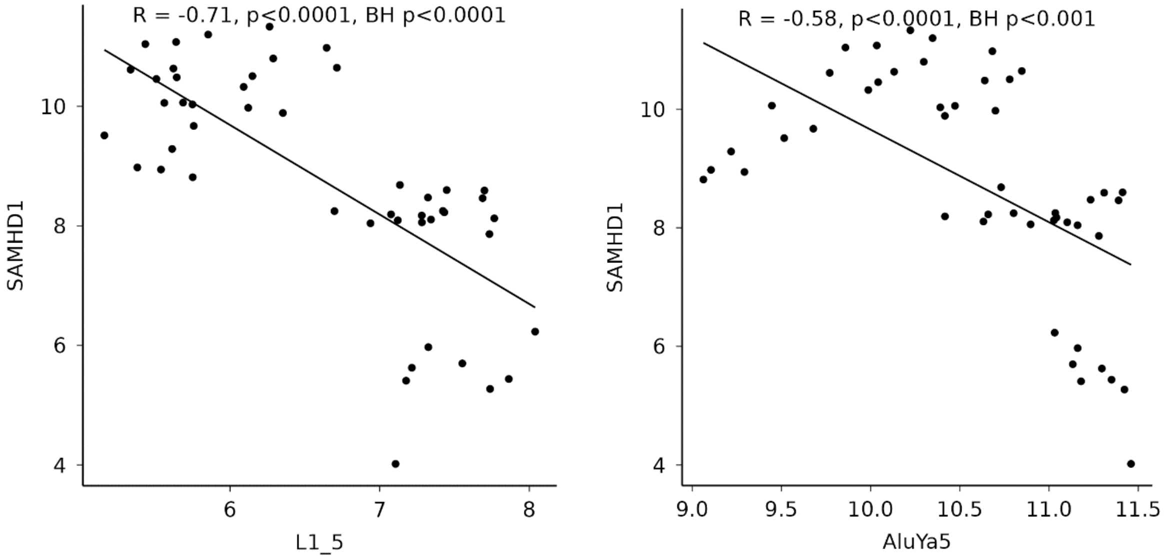


Gene expression correlation between *SAMHD1* and LINE1 (L1_5) and Alu (AluYa5) in peripheral blood pooled circulating lymphocytes subsets. R: Pearson's correlation coefficient.

**Supplementary file 13**

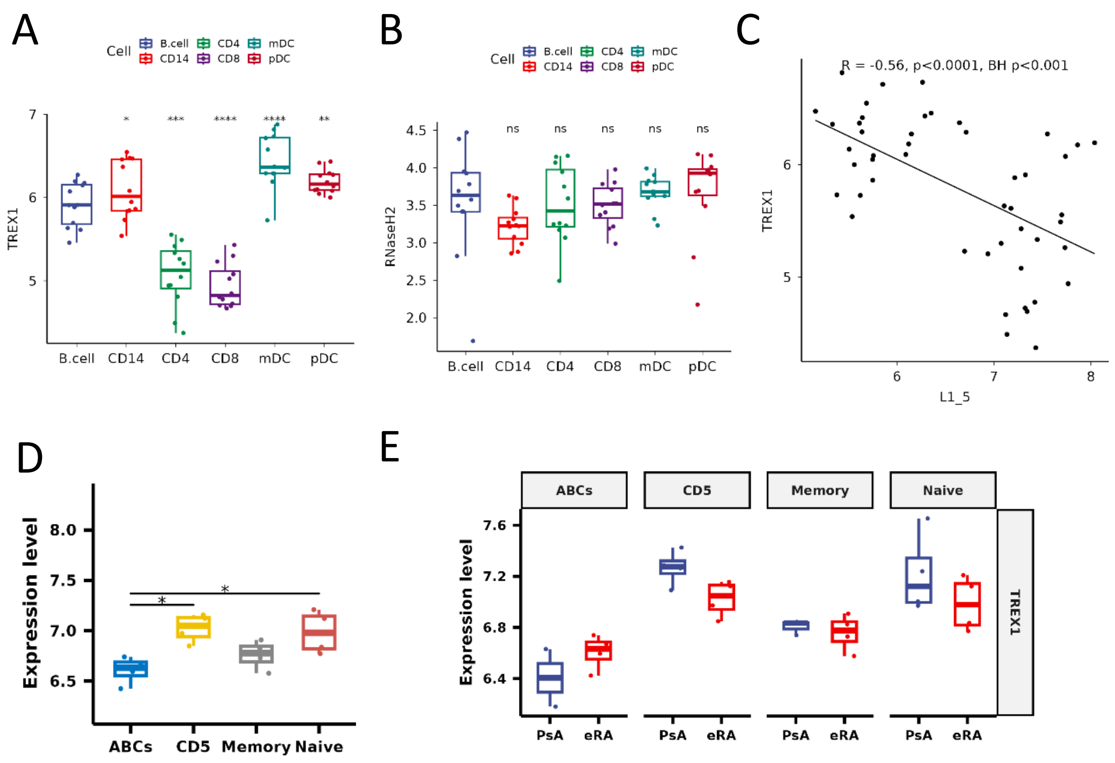


**(A)** Expression of *TREX1* and **(B)** *RNAseH2* as determined by Nanostring Technology in flow cytometry sorted peripheral blood lymphocyte subsets (plasmacytoid dendritic cells [pDCs], conventional CD1c$^{+}$ DCs [mDCs], CD4$^{+}$ T cells (CD4), CD8$^{+}$ T cells (CD8), whole CD19$^{+}$ B cells, and CD14$^{+}$ monocytes (CD14) from early RA patients (eRA, n=8), B cells used as a reference in pairwise paired t tests. **(C)** Pearson's correlation coefficient of *TREX1* and LINE 1 (L1 5'UTR) in from these same circulating lymphocytes pooled, BH corrected p<0.0001 **(D)** Flow cytometrically naïve B cells (CD19$^{+}$IgD$^{+}$CD27$^{-}$), memory B cells (CD19$^{+}$IgD$^{-}$CD27$^{+}$), age associated B-cells (ABCs, CD19$^{+}$CD11c$^{+}$CD21$^{-}$) and CD5$^{+}$ B cells (CD19$^{+}$CD5$^{+}$) were examined from both early RA patients (eRA) and disease control, early psoriatic arthritis patients (PsA) and differences in *TREX1* expression, determined by Nanostring Technologies, was examined between early RA specific B-cell subsets and **(E)** disease cohorts, Wald test with adjusted and raw P values shown for 3D and 3E respectively.  ***p<0.001

**Supplementary file 14**

| Class | Number of EREs | cell | Fold_Change direction |
|---|---|---|---|
| LINE | 13 | B Cell | Increased in RA |
| LTR | 219 | B Cell | Increased in RA |
| SINE | 1 | B Cell | Increased in RA |
| Other | 80 | B Cell | Increased in RA |
| LINE | 133 | B Cell | Decreased in RA |
| LTR | 98 | B Cell | Decreased in RA |
| SINE | 52 | B Cell | Decreased in RA |
| Other | 149 | B Cell | Decreased in RA |
| LINE | 12 | Fibroblast | Increased in RA |
| LTR | 211 | Fibroblast | Increased in RA |
| Other | 76 | Fibroblast | Increased in RA |
| LINE | 120 | Fibroblast | Decreased in RA |
| LTR | 131 | Fibroblast | Decreased in RA |
| SINE | 58 | Fibroblast | Decreased in RA |
| Other | 166 | Fibroblast | Decreased in RA |
| LINE | 24 | Monocyte | Increased in RA |
| LTR | 317 | Monocyte | Increased in RA |
| SINE | 1 | Monocyte | Increased in RA |
| Other | 112 | Monocyte | Increased in RA |
| LINE | 51 | Monocyte | Decreased in RA |
| LTR | 23 | Monocyte | Decreased in RA |
| SINE | 51 | Monocyte | Decreased in RA |
| Other | 72 | Monocyte | Decreased in RA |
| LINE | 14 | T Cell | Increased in RA |
| LTR | 192 | T Cell | Increased in RA |
| Other | 76 | T Cell | Increased in RA |
| LINE | 107 | T Cell | Decreased in RA |
| LTR | 99 | T Cell | Decreased in RA |
| SINE | 54 | T Cell | Decreased in RA |
| Other | 141 | T Cell | Decreased in RA |

In single cell sequencing analysis of established rheumatoid arthritis (RA) synovial tissue compared with osteoarthritis synovial tissue the number of individual endogenous retroelements (ERE) that have either increased or decreased expression in each cellular subset, as shown in figure 5B.

**Supplementary file 15**

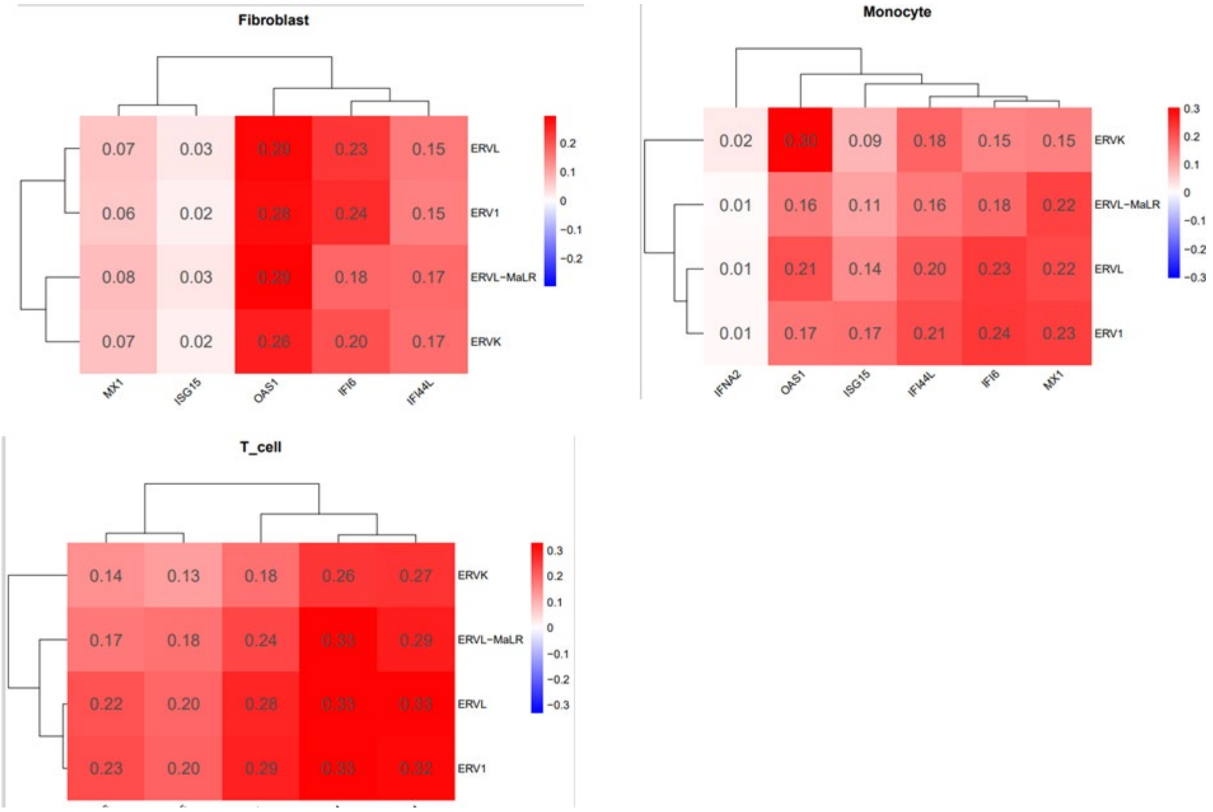


Hierarchical clustering of correlations between gene expression of interferon response genes and endogenous retroelement expression counts from single cell RNA seqencing data from established RA synovium. Pearson's correlation co-efficients depicted. All correlations were significant, p<0.001.
